# Population level consumption of cephalosporins and macrolides may select for reduced antimicrobial susceptibility to unrelated antimicrobials in *Neisseria gonorrhoeae*: an ecological analysis

**DOI:** 10.1101/2020.07.08.20148957

**Authors:** Chris Kenyon

**Affiliations:** Professor in Sexually Transmitted Infections HIV/STI Unit, Institute of Tropical Medicine, Antwerp, Belgium; Division of Infectious Diseases and HIV Medicine, University of Cape Town, Anzio Road, Observatory 7700, South Africa

## Abstract

**Aims:** Previous studies have found positive associations between population level consumption of macrolides/cephalosporins/floroquinolones and *Neisseria gonorrhoeae* sensitivity to the same class of antimicrobial. We hypothesized that these associations may also apply to different classes of antimicrobials.

**Methods:** Using susceptibility data from 24 countries in the European Gonococcal Antimicrobial Surveillance Programme and antimicrobial consumption data from the IMS Health MIDAS database we built mixed effects linear regression models with country-level cephalosporin, quinolone and macrolide consumption (standard doses/ 1000 population/ year) as the explanatory variables (from 2009 to 2015) and 1-year lagged ceftriaxone, azithromycin and ciprofloxacin geometric mean minimum inhibitory concentrations and percent resistance as the outcome variables (2010 to 2016).

**Results:** Positive associations were found between the consumption of cephalosporins and geometric mean MIC ceftriaxone, cefixime and ciprofloxacin. Macrolide consumption was associated with cefixime and ciprofloxacin geometric mean MIC as well as the prevalence of antimicrobial resistance to ciprofloxacin and cefixime.

**Conclusions:** Population level consumption of cephalosporins and macrolides may select for reduced antimicrobial susceptibility to unrelated antimicrobials.

## Background

*Neisseria gonorrhoeae* (Ng) has acquired antimicrobial resistance to most classes of antimicrobials, including combined high level resistance to ceftriaxone and azithromycin [1, 2]. Both in vitro and individual level studies have established a link between antimicrobial exposure and the development of antimicrobial resistance (AMR) in Ng [3-10]. Antimicrobials used to treat Ng (and other STIs) are one important determinant of AMR in Ng [11, 12].

The consumption of antimicrobials in the general population (bystander selection) is another determinant. A study of all countries contributing data to the World Health Organizations Global Gonococcal Antimicrobial Surveillance Programme found positive associations between the consumption and resistance for cephalosporins, macrolides and quinolones [13]. A previous ecological study of 24 countries in Europe found that the population-level consumption of cephalosporins was associated with the Ng geometric mean MIC of ceftriaxone and cefixime [14]. Likewise, fluoroquinlone consumption was positively associated with the prevalence of fluoroquinolone resistance.

Using the same dataset as this previous study, we aimed to test the hypothesis that the consumption of antimicrobials in the general population induces AMR in Ng to unrelated antimicrobials. A number of in vitro and epidemiological studies have revealed this effect for a range of other bacterial species [15-18]. A case control study of risk factors associated with Methicillin Resistant Staphylococcus Aureus (MRSA) infection, for example, found that exposure to any antibiotic in the preceding year was associated with an approximate tripling of the risk of MRSA [19]. This effect was strongest for quinolones and macrolides, rather than β-lactams. The activation of multidrug efflux pumps plays an important role in the induction on non-homologous resistance. β-lactams can, for example, select for resistance to quinolones (and vice versa) in *Pseudomonas aeruginosa* via this mechanism [17]. A detailed in vitro study of the pathways of cross-resistance in *Escherichia coli*, revealed that single antimicrobial exposure led to multidrug resistance in the majority of antimicrobial pairs tested [20]. Of relevance to our study, macrolides, quinolones and cephalosporins were all found to induce AMR to one another. At a population level, studies from Spain, have found that macrolides were more important drivers for local differences in both erythromycin and penicillin resistance in *Streptococcus pneumoniae* than β-lactams were [15, 16]. A multi-country European study confirmed these findings and in addition found associations between quinolone consumption and erythromycin and penicillin resistance in *S. pneumoniae* [21]. Whilst epidemiological studies have not established such nonhomologous effects in Ng, various considerations make this a worthwhile hypothesis to test. One study found that inactivating the MtrCDE multidrug efflux pump in Ng (which can be selected for by cephalosporins and macrolides [22]) results in large reductions in minimum inhibitory concentrations (MICs) to ciprofloxacin (a quinolone), ceftriaxone (a cephalosporin) and azithromycin (a macrolide) [23]. Furthermore a number of studies have found positive associations between AMR to different antimicrobials in isolates of Ng [6, 24, 25].

## Methods

Our data came from two sources.

### Antimicrobial resistance

Euro-GASP includes a sentinel surveillance programme that tests a representative number of isolates from 28 European Union/European Economic Area member states per year for a range of antimicrobials through a biannual hybrid centralised/decentralised system (Table 1). We used the Euro-GASP MIC data for azithromycin, cefixime, ceftriaxone and ciprofloxacin between 2010 and 2016. The full Euro-GASP methodology, including suggested sampling strategy, laboratory techniques, external quality assurance and internal quality control mechanisms has been published elsewhere [26]. The following minimum inhibitory concentration (MIC) breakpoints are used to define antimicrobial resistance in EuroGASP: Azithromycin: >0.5 mg/L, Cefixime: >0.12 mg/L, Ceftriaxone: >0.12 mg/L, Ciprofloxacin: >0.06 mg/L [27, 28].

**Table 1.** List of countries included in study and method used to assess antimicrobial sensitivity

| Country | Country code | Method of testing |  |
| --- | --- | --- | --- |
| Austria | AT | Decentralised | Etest |
| Belgium | BE | Decentralised | AD |
| Croatia | HR | Centralised | Etest |
| Czech Rep. | CZ | Decentralised | Etest |
| Denmark | DE | Decentralised | Etest |
| Estonia | EE | Centralised | Etest |
| France | FR | Decentralised | Etest |
| Germany | DE | Centralised | BKP/Etest |
| Greece | EL | Decentralised | Etest |
| Hungary | HU | Centralised | BKP/Etest |
| Iceland | IS | Decentralised | Etest |
| Ireland | IE | Decentralised | Etest |
| Italy | IT | Decentralised | Etest |
| Latvia | LV | Centralised | Etest |
| Finland | FI | Decentralised | Etest |
| Netherlands | NL | Decentralised | Etest |
| Norway | NO | Decentralised | AD |
| Poland | PL | Centralised | Etest |
| Portugal | PT | Decentralised | Etest |
| Slovakia | SK | Centralised | Etest |
| Slovenia | SI | Decentralised | Etest |
| Spain | ES | Decentralised | Etest |
| Sweden | SE | Decentralised | Etest |
| UK | UK | Decentralised | AD/Etest |

Etest minimum inhibitory concentration (MIC) gradient strips to determine the MIC of an antimicrobial, AD agar dilution method, BKP Breakpoint agar dilution method

### Antimicrobial consumption

Data from Intercontinental Marketing Statistics Health MIDAS (IMS Health, Danbury, CT, USA) were used as a measure of national antimicrobial drug consumption. IMS uses national sample surveys that are performed by pharmaceutical sales distribution channels to estimate antimicrobial consumption from the volume of antibiotics sold in retail and hospital pharmacies. The sales estimates from this sample are projected with use of an algorithm developed by IMS Health to approximate total volumes for sales and consumption. Antimicrobial consumption estimates are reported as the number of standard doses (a dose is classified as a pill, capsule, or ampoule) per 1000 population per year. Data on consumption of cephalosporins, quinolones and macrolides was available for 34 European countries. We used data for the 24 countries that were also represented in the Euro-GASP dataset.

### Data analysis

For each antimicrobial we calculated the following variables by country and year: 1) Geometric mean MIC of all samples, 2) the percentage of isolates with antimicrobial resistance.

The association between the sensitivities (AMR and MIC) of different antimicrobials at an individual and country level was assessed using mixed effects logistic regression. The MIC values were log transformed to create more normal distributions for these regression analyses.

For each antimicrobial, mixed effects linear regression was used to assess the association between antimicrobial consumption and (1) geometric mean MIC and (2) the percentage of isolates with antimicrobial resistance. All country level mixed effects analyses were weighted by the number of samples used in the analysis.

The statistical analyses were performed in R V.3.3.2 and Stata 13.0.

## Results

The number of countries with antimicrobial consumption data reported increased from 19 countries in 2009 to 24 countries in 2015 (Table 1). Likewise, the completeness of data for AMR improved by year for all antimicrobials (details reported in [14]). Large variations in antimicrobial consumption and AMR were evident between countries. These have been reported elsewhere [14, 21].

### Associations between sensitivities of different antimicrobials

#### Individual isolate level

AMR for all four antimicrobials assessed were statistically significantly positively associated with one another (coef. from 0.15 (95% confidence interval [CI] 0.07-0.22) to 2.2 (95% CI 1.9-2.6) all P <0.05; Table 2). The same was true for the log transformed MIC values (coef. from 0.05 to 2.3; all P <0.005; Table 2).

**Table 2.** Adjusted individual isolate level associations: A) between antimicrobial resistance in azithromycin, cefixime, ceftriaxone and ciprofloxacin [coefficients (95% Confidence Intervals) reported; assessed via mixed effects logistic regression]; B) between log transformed geometric mean MIC for azithromycin, cefixime, ceftriaxone and ciprofloxacin [coefficients (95% Confidence Intervals) reported; assessed via mixed effects linear regression] ^a.^ In all the calculations, the row variable is the outcome variable.

| Individual isolate level |  |  |  |  |
| --- | --- | --- | --- | --- |
|  | Azithromycin | Cefixime | Ceftriaxone | Ciprofloxacin |
| **A) Antimicrobial Resistance** |  |  |  |  |
| Azithromycin | - | 0.83 (0.71-0.94)** | 0.61 (0.10-1.12)* | 0.15 (0.07-0.22)** |
| Cefixime | 0.74 (0.63-0.86)** | - | 2.0 (1.5-2.5)** | 2.1 (1.8-2.5)** |
| Ceftriaxone | 0.60 (0.09-1.10)* | 2.1 (1.6-2.7)** | - | <sup>b</sup> |
| Ciprofloxacin | 0.17 (0.010-0.24)** | 2.24 (1.87-2.62)** | <sup>b</sup> | - |
| **B) MIC** |  |  |  |  |
| Azithromycin | - | 0.37 (0.34-0.39)** | 0.32 (0.31-0.34)** | 0.05 (0.05-0.06)** |
| Cefixime | 0.22 (0.21-0.23)** | - | 0.54 (0.53-0.54)** | 0.11 (0.10-0.11)** |
| Ceftriaxone | 0.32 (0.30-0.34)** | 0.90 (0.88-0.91)** | - | 0.17 (0.17-0.18)** |
| Ciprofloxacin | 0.69 (0.62-0.75)** | 2.3 (2.3-2.3)** | 2.2 (2.2-2.3)** | - |

<sup>a</sup> Adjusted for year of isolate, random intercept for country
\*P<0.05 \*\*P<0.005
<sup>b</sup> All 26 isolates with ceftriaxone resistance had ciprofloxacin resistance and thus the model did not achieve convergence

##### Country level

Azithromycin resistance was a significant predictor of resistance to cefixime (coef. 0.18; P <0.05) and ciprofloxacin (coef. 0.31; P < 0.05; Table 3), as was cefixime resistance a predictor for resistance to azithromycin (coef. 5.7; P <0.005), ceftriaxone (coef. 0.03; P <0.05) and ciprofloxacin (coef. 0.79; P <0.005). Ceftriaxone resistance predicted resistance to azithromycin (coef. 6.7; P < 0.005) and cefixime (coef. 1.5; P < 0.05), whereas ciprofloxacin resistance predicted resistance to cefixime (coef. 0.18; P < 0.005) and ceftriaxone (coef. 1.5; P < 0.05). The geometric mean MICs of the four antimicrobials were all significantly positively associated with one another excluding the cefixime-ciprofloxacin comparison (coef. 0.003 to 58.3; all P < 0.05).

**Table 3.**
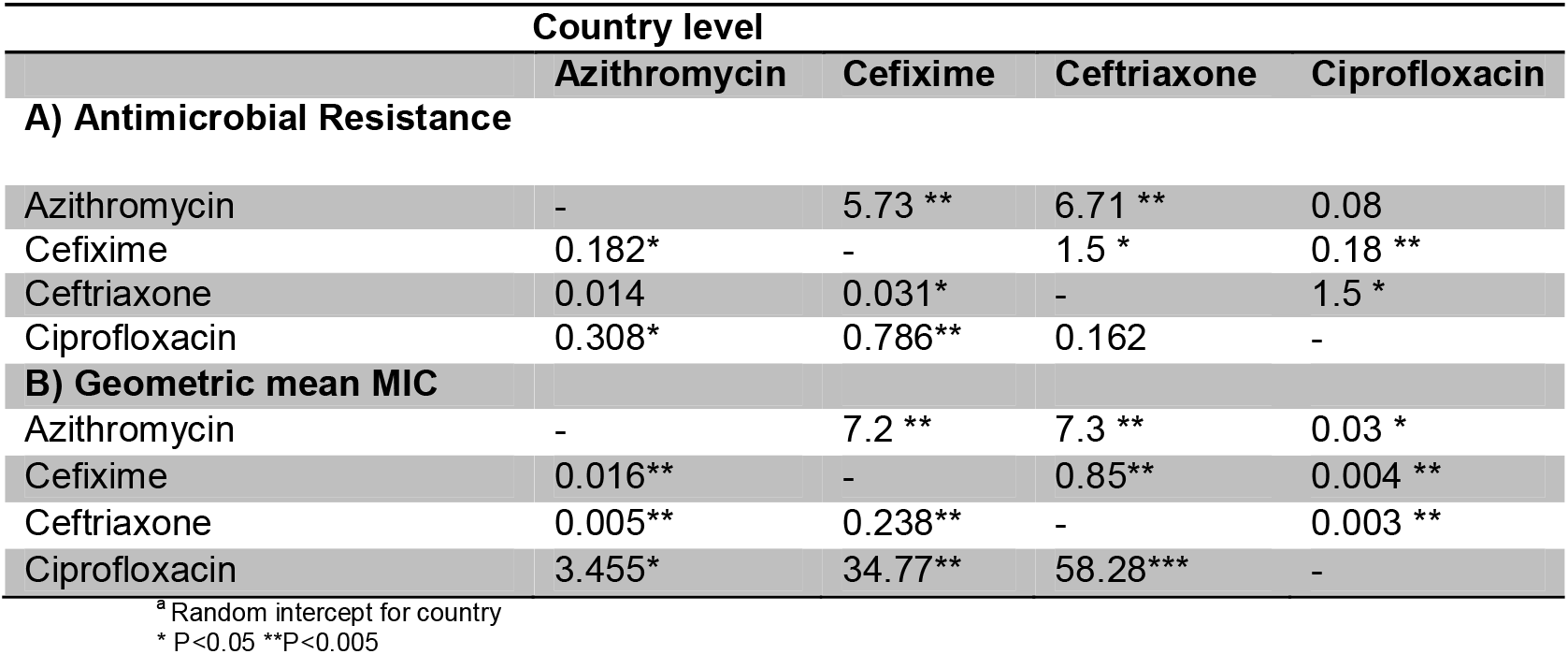
Country level associations: Mixed effects linear regression of A) percent antimicrobial resistance and B) geometric mean MIC of azithromycin, cefixime, ceftriaxone and ciprofloxacin given the value for the percent resistance/ geometric mean MIC of the other antimicrobial (coefficients). In all cases the row variable is the outcome variable ^a^

### Correlation between antimicrobial sensitivity and consumption

Linear regression analyses revealed positive associations between the consumption of cephalosporins and geometric mean MIC ceftriaxone, cefixime and ciprofloxacin (all P’s < 0.05; Table 4). Macrolide consumption was associated with cefixime and ciprofloxacin geometric mean MIC (all P’s < 0.05). Rerunning the models with AMR as the independent variable, macrolide consumption was associated with cefixime resistance (Table 4; P = 0.038) and cephalosporin, macrolide and quinolone consumption were all associated with ciprofloxacin resistance (all P < 0.01).

**Table 4.** Mixed effects linear regression analyses of relationship between antimicrobial consumption (doses/population) and geometric mean MIC/ antimicrobial resistance

| Dependent variable | Independent variable | Geometric mean MIC |  | Antimicrobial resistance |  |
| --- | --- | --- | --- | --- | --- |
|  |  | Coefficient | P-value | Coefficient | P-value |
| **Y/MIC<sub>CRO</sub>** | C<sub>Cephalosporin</sub> | 0.003 | 0.003 | 0.2 | 0.073 |
|  | C<sub>Macrolide</sub> | 0.004 | 0.072 | 0.1 | 0.607 |
|  | C<sub>Quinolone</sub> | 0.001 | 0.068 | 0.2 | 0.377 |
| **Y/MIC<sub>CFM</sub>** | C<sub>Cephalosporin</sub> | 0.004 | 0.017 | 1.6 | 0.062 |
|  | C<sub>Macrolide</sub> | 0.005 | 0.014 | 2.3 | 0.038 |
|  | C<sub>Quinolone</sub> | 0.007 | 0.071 | 1.9 | 0.137 |
| **Y/MIC<sub>AZM</sub>** | C<sub>Cephalosporin</sub> | 0.04 | 0.115 | 1.7 | 0.257 |
|  | C<sub>Macrolide</sub> | 0.01 | 0.497 | 3.0 | 0.086 |
|  | C<sub>Quinolone</sub> | -0.02 | 0.640 | -0.7 | 0.835 |
| **Y/MIC<sub>CIP</sub>** | C<sub>Cephalosporin</sub> | 0.46 | <0.0001 | 2.7 | 0.001 |
|  | C<sub>Macrolide</sub> | 0.51 | 0.0001 | 13.2 | <0.001 |
|  | C<sub>Quinolone</sub> | 0.49 | 0.063 | 16.8 | 0.006 |

\* P<0.05; \*\* P<0.005; \*\*\* P<0.0005; CRO – ceftriaxone, CFM – cefixime, AZM – azithromycin, CIP - Ciprofloxacin

## Discussion

Building on a previous study that found associations between population level consumption of cephalosporins and fluoroquinolones and homologous class susceptibility, we found positive associations between non-homologous class associations. These associations were present at the level of both individual isolates and countries. At an individual level both MICs and antimicrobial resistance for azithromycin, cefixime, ceftriaxone and ciprofloxacin were all positively associated with one another. With a few exceptions, the same was true for geometric mean MIC and the prevalence of antimicrobial resistance at a country level.

We also found positive associations between country-level geometric mean MIC/AMR and of non-homologous class antimicrobial consumption. This effect was most noticeable for macrolides. For example, macrolide consumption was a stronger predictor of ciprofloxacin/cefixime geometric MIC than fluoroquinolone/cephalosporin consumption, respectively.

These findings are commensurate with those from other bacteria where similar associations have been found [21, 29-34]. Both macrolides and quinolones have been found to be associated with β-lactam resistance in other organisms but this is the first time that we are aware of that this effect has been demonstrated in Ng [15-17, 35, 36]. The fact that positive associations between consumption and AMR/MIC were also found at an individual isolate level between all pairs of antimicrobials tested makes these findings less likely to be due to the ecological inference fallacy. Further work is required to test in vitro and at individual and population levels if differential effects of macrolides versus β-lactams on resistant versus sensitive subpopulations of Ng may play a role in macrolides induction of resistance as has been observed for other bacteria [37-39].

There are however a number of other explanations for these findings. The consumption of the three classes of antimicrobials were themselves positively associated with one another which means the positive country level associations we found between non-homologous consumption and reduced susceptibility may be explained by confounding. We did not control for this at country level as it is unclear to us if this would be meaningful in this type of ecological analysis. There were also a large number of other factors responsible for the emergence of AMR which we did not control for [11, 12, 40]. Further study limitations include incomplete data and/or small samples for a number of countries.

If this link between antimicrobial consumption and resistance applies to Ng as well as other bacteria, then this would have important public health consequences. Baquero et al., have argued persuasively that the linked emergence of Extended Spectrum Beta Lactamase (ESBL) resistance in multiple bacterial species in hospitals in Spain was best viewed as a syndemic rather than epidemics of single species or clones [41]. Appreciating this as a syndemic enabled them to address the underlying environmental determinant of AMR: excess use of antimicrobial drugs that induce ESBL production in multiple bacteria species rather than traditional approaches targeting individual clones or species [41, 42]. Likewise, efforts to reduce antimicrobials particularly associated with the genesis of antimicrobial resistance such as macrolides, cephalosporins and fluoroquinolones in Scotland has been shown to have resulted in a reduction in the prevalence of methicillin resistance of *Staphylococcus aureus* [43].

Whilst further in-vitro and clinic-epidemiological studies are required to establish the extent to which cross resistance emerges in Ng our study results provide further motivation to reduce the consumption of antimicrobials, such as macrolides, as far as is possible and safe in the general population.

## Data Availability

Data is available as detailed in the manuscript

## Acknowledgements

Data from The European Surveillance System – TESSy, provided by countries listed in Table 1 and released by ECDC

## Consent for publication

Not applicable

## Disclaimer

The views and opinions of the authors expressed herein do not necessarily state or reflect those of ECDC. The accuracy of the authors’ statistical analysis and the findings they report are not the responsibility of ECDC. ECDC is not responsible for conclusions or opinions drawn from the data provided. ECDC is not responsible for the correctness of the data and for data management, data merging and data collation after provision of the data. ECDC shall not be held liable for improper or incorrect use of the data.

